# “We also deserve help during the pandemic”: The effect of the COVID-19 pandemic on foreign domestic workers in Hong Kong

**DOI:** 10.1101/2021.03.04.21252889

**Authors:** Ingrid D. Lui, Nimisha Vandan, Sara E. Davies, Sophie Harman, Rosemary Morgan, Julia Smith, Clare Wenham, Karen Ann Grépin

## Abstract

The coronavirus disease 2019 (COVID-19) pandemic poses particular challenges for migrant workers around the world. This study explores the unique experiences of foreign domestic workers (FDWs) in Hong Kong, and how COVID-19 impacted their health and economic wellbeing. Interviews with FDWs (*n* = 15) and key informants (*n* = 3) were conducted between May and August 2020. FDWs reported a dual-country experience of the pandemic, where they expressed concerns about local transmission risks as well as worries about their family members in their home country. Changes to their current work situation included how their employers treated them, as well as their employment status. FDWs also cited blind spots in the Hong Kong policy response that also affected their experience of the pandemic, including a lack of support from the Hong Kong government. Additional support is needed to mitigate the particularly negative effects of the pandemic on FDWs.

## 1. Introduction

Migrants are particularly vulnerable to the direct and indirect effects of the coronavirus disease 2019 (COVID-19) pandemic. The International Organization for Migration (IOM) has found that migrants were often excluded from host country welfare systems, were greatly affected by border closures which limited their ability to travel between their home and host countries, and that the pandemic was used as an opportunity to foster xenophobic and discriminatory attitudes towards them [1]. Migrants also often face additional barriers in accessing healthcare or information about COVID-19 in their host countries, increasing their risk of contracting the disease [2]. The financial impact of the pandemic has been forecasted to result in a $108.6 billion decline in global remittances, assuming recovery within a year, putting remittance-dependent households at risk of falling into poverty [3].

Foreign domestic workers (FDWs) are migrants who engage in work performed in or for a household within an employment relationship [4]. In 2015, it was estimated that there were 11.5 million FDWs in the world, with 73.4% of them being women [5]. By outsourcing domestic work, more married women have been able to enter full-time paid work, creating dual-earner households with a higher household income, benefitting the host country economy [6]. Women tend to migrate to work in domestic and caretaking roles, whereas men typically take up jobs in sectors like construction, which is significant as these different occupations are likely to have their own health risks [7], affecting their susceptibility to COVID-19.

The lack of recognition and supportive policies for migrants can have many physical, mental, social, and economic effects on FDWs. Live-in FDWs tend to work long hours with an unspecified workload while living in the same place that they work in, making it difficult to mentally separate work from private time [8]. They are also vulnerable to abuse from their employers due to the unequal power relationship, including verbal abuse and physical assault, and are often unable to negotiate for or assert their rights [9]. FDWs often also struggle with the relationships with their families back home. For example, FDWs may perceive that their relatives only value the remittances they send, devaluing the work and sacrifices they are making overseas [10]. As FDWs are often visible minorities within their host country, they may also face discrimination as migrants, minorities, and women [7].

In Hong Kong, FDWs are essential to the functioning of many local families, but they are also regarded as separate from the “real family” [6]. While they make significant contributions to Hong Kong’s economy, they are still denied economic rights [11]. In 2019, the Immigration Department reported that there were 399,320 FDWs in Hong Kong, with 98.5% of them being women, and most of them coming from the Philippines (55%), Indonesia (43%), or Thailand (0.5%) [12]. Given the enormous health, economic, and social effects of the COVID-19 pandemic, we sought to explore the effects of the pandemic on FDWs in Hong Kong through in-depth interviews with FDWs and key informants to better understand their experiences. We focused on how this minority population, comprised mostly of women, was disproportionately affected by the pandemic response in terms of their health and economic wellbeing. A greater understanding of the experiences of this vulnerable population can help to guide the policies aimed at mitigating the negative effects of the pandemic on society in Hong Kong and in other areas where there are significant FDW populations.

### 1.1 The status of FDWs in Hong Kong

According to Hong Kong laws, an FDW’s ability to stay in Hong Kong is bound by a renewable two-year Standard Employment Contract [13], which states that they can only provide full-time, live-in domestic services (including household chores, cooking, baby-sitting, child-minding, and looking after aged persons) at a specific employer’s residence. The Contract also outlines the obligations of the employer. First, employers are required to pay the FDW no less than the Minimum Allowable Wage, which, as of September 2019, was $4,630 HKD (∼$600 USD) per month [14]. They are also required to provide FDWs with suitable accommodation within their residence as well as free food or a food allowance of no less than $1,121 HKD ($145 USD) per month. FDWs are entitled to at least one rest day per week, usually on Sundays, paid statutory holidays, and paid annual leave, as well as a vacation to their home country at the end of each two-year contract period. Finally, either party may terminate the contract with fair notice, barring extenuating circumstances, such as if an employer believes the FDW has neglected their duties, or if an FDW is subject to ill-treatment by the employer [15]. On top of the Contract, FDWs enjoy the same legal protections under the Employment Ordinance as other employees in Hong Kong, including a right to form labour unions. They are also supported by numerous local NGOs that have worked to amplify their voices and advocate for their rights [9]. FDWs who experience abuse or exploitation have several options for recourse, including calling the police, contacting the Labour Department or their government’s Consulate-General in Hong Kong, and submitting a complaint to the Equal Opportunities Commission for discrimination-related cases.

Despite these protections, FDWs still face a number of disadvantages. According to the Immigration Ordinance, a person who has resided in Hong Kong for a continuous period of not less than seven years may apply for permanent residency, but FDWs are explicitly exempted from this regardless of how long they have remained in the city [16]. They have also been prohibited by law from living outside of their employer’s residence since 2003, a ruling which was recently upheld again in September 2020 [17]. However, the average size of a Hong Kong apartment is relatively small, and there is no clear definition what constitutes a “suitable” accommodation. One study from a local charity, Mission for Migrant Workers, investigated the living accommodations of FDWs, and found that three out of five FDWs they studied either did not have their own room to sleep in, or their room was also utilized for other purposes, such as a storage area for the household. While some of those who did not have their own room shared a bedroom with someone else (e.g., children), others reported sleeping in the living room, the kitchen, or even the bathroom [18].

Poor living conditions of this nature have been linked to poor mental health outcomes: one study reported that FDWs who were less satisfied with their living accommodations were also more likely to have higher levels of depression [19]. Studies have also shown that FDWs tend to have lower levels of self-reported physical and mental health than the general adult population of Hong Kong, with factors such as a larger household size and increased working hours contributing to poorer physical health [20]. Factors such as withheld wages, the inability to regularly send remittances back home, physical abuse and discrimination contributed to poorer mental health, while daily contact with friends was associated with better mental health [20].

Furthermore, when an FDW’s contract has been terminated, they are required to leave Hong Kong within two weeks from the date of termination, unless they find a new employer. If they overstay in Hong Kong, they could be prosecuted and subjected to a fine of up to $50,000 HKD ($6,450 USD), and imprisonment for up to two years. After serving their sentence, they will be deported from Hong Kong and will not be permitted to work in Hong Kong as FDWs again [15]. This is significant because it typically takes four to six weeks for a change of employment to be processed, leading many FDWs to stay with abusive employers due to a fear of losing their job [19]. Many instances of abuse against FDWs have been reported in Hong Kong, with one study demonstrating that they are vulnerable to sexual abuse, as well as psychological abuse [21]. Many survivors remained working for the abusive employer, while a few chose to quit and many others had difficulty deciding whether to stay or quit [21]. Abuse is used by employers to display power, reinforce the employer-employee hierarchy and dehumanize FDWs [9].

Race and ethnicity also play a key role, intersecting with gender to further marginalize FDWs in Hong Kong. There are specific stereotypes about different ethnicities of FDWs: Filipinos are seen as more highly educated with better English skills [22], whereas Indonesians are viewed as requiring more micro-management by employers [23]. These stereotypes are further supported by the Hong Kong press, which consistently portrays an “us-them” dichotomy between employers and FDWs, reinforcing cultural perceptions of FDWs as “alien” or “other” and legitimising the exploitation and abuse they face [24].

### 1.2 The COVID-19 pandemic in Hong Kong

The Hong Kong government began to respond to the pandemic in early January 2020, days after a cluster of viral pneumonia with unknown aetiology was first detected in Wuhan, China [25]. This included a series of travel restrictions mainly aimed at travellers from mainland China, however, measures escalated into the lead up to the Chinese New Year holiday period in late January, which is a time where many local families normally travel abroad and many FDWs take their annual leave. On February 2, the Philippine President Duterte issued a temporary ban on Filipinos from travelling to Hong Kong, mainland China and Macau [26], leaving many FDWs who had gone home over the holidays stranded, and their employment contracts with their employers uncertain [27]. FDWs were exempted from the outbound travel ban two weeks later on February 18 [28], but restrictions on inbound travellers were imposed by the Hong Kong government on March 18, including a compulsory two-week quarantine for all inbound travellers [29]. In July, several high-risk countries were added to a list of specified places wherein travellers from these places had to present a valid negative COVID-19 test result and quarantine in a hotel upon arrival, including Bangladesh, India, Indonesia, Nepal, Pakistan, and the Philippines [30]. By July, employers were required to bear the cost of the nucleic acid test, as well as provide accommodation and food expenses for FDWs during their compulsory quarantine [31].

However, despite the government regulations, many families were not keen to have an FDW from abroad carry out their quarantine in the family home when such measures had been permitted, nor pay for the additional expenses once additional measures had been imposed. Some FDWs stayed in temporary boarding houses to undertake their quarantines. Such houses were also being used by FDWs who were between contracts and were trying to secure a new contract or had to return to their home countries. These facilities are usually small, and during the pandemic up to 20 FDWs could be living in a 400 sq ft apartment at once [32]. Perhaps unsurprisingly, two COVID-19 clusters have been linked to such facilities, first in August and then December, 2020.

While initially the Hong Kong response was lauded for being both quick and effective, this high level perspective overlooks the many challenges the government faced in launching its response, including the fact that the region had just undergone an extended period of civil unrest and prevailing high levels of mistrust in government [33, 34]. Indeed, the actions of Hong Kong society in response to COVID-19 stemmed from the memories of the SARS outbreak in 2003 and dissatisfaction with the government, leading to faster adoption of personal hygiene measures and networks of community-based assistance [35]. Their policies have also largely lacked support and protection for vulnerable groups, including FDWs.

## 2. Material and methods

### 2.1 Participants

This study is based on data collected from interviews with 15 FDWs who worked in Hong Kong during the COVID-19 pandemic. FDWs were recruited as part of a larger project that aimed to understand and mitigate the real-time differential gendered effects of the COVID-19 outbreak^1^. All participants were women: ten participants were from the Philippines, one was from Indonesia, one was from Thailand, and three were from Sri Lanka. Purposive convenience sampling was employed to recruit FDWs through social media adverts. A small incentive gift worth $50 HKD (∼$6 USD) in the form of a gift certificate for a popular coffee shop was provided to all of the FDW participants. This study also included three key informants from organizations that work closely with FDWs in Hong Kong, who were recruited purposefully in accordance with their professional roles in current decision-making bodies related to outbreak response and gender equity initiatives. No individual incentives were provided to the key informants. All participants have been assigned numeric codes (e.g., FDW1 and KI1 for FDW participants and key informants, respectively) to ensure the anonymity of our respondents.

### 2.2 Materials and procedure

A semi-structured interview guide was developed, which included a total of 6 questions for the FDWs and 8 questions for the key informants. This guide was based on the domains of the COVID-19 Gender Matrix, which takes a multidimensional perspective on the effects of COVID-19 [36]. Probing follow-up questions were used to further understand participants’ experiences.

The interviews lasted an average of 31 minutes. All interviews were conducted between May and August 2020 and were done over the phone or Zoom. Each interview was audio-recorded with verbal consent from each participant, and notes were taken during each interview. All interviews were carried out by one researcher and were conducted in English. All recordings were then securely stored on the online Simon Fraser University Vault. Interviews were transcribed verbatim and checked against audio recordings for accuracy. Ethics approval was obtained from the Human Research Ethics Committee of the University of Hong Kong.

### 2.3 Analysis

Interviews were analysed using the framework approach as outlined by Smith and Firth [37]. Two researchers read through all transcripts and created an initial coding guide based on recurring themes that arose from the data. The two sets of codes were then compared and synthesised into one coding index. One researcher subsequently coded all transcripts using the coding index, which was refined through ongoing analysis, including the addition of new categories and the grouping of similar categories into larger themes. The coding index was subjected to second-order analysis to narrow down themes that were specific to FDWs. Second-order analysis was undertaken to determine final themes which emerged most frequently across all FDW participants and key informants.

## 3. Results

FDWs faced unique challenges that affected their experiences during the COVID-19 pandemic. Below we present key themes from the pandemic in Hong Kong that we identified as specific to FDWs.

### 3.1 Dual-country experience of the pandemic

FDWs were concerned both about the pandemic in Hong Kong as well as the effect it was having directly and indirectly on their families and home countries. This was demonstrated by quotes from FDW11, who was worried about the risk of infection in Hong Kong with the effects it was likely to have on her family back home: *“If I get this virus, what will happen to me? What will happen to my family?”* FDWs were also very concerned about the state of the pandemic back in their home countries and the lack of control of the outbreak relative to the experience in Hong Kong, especially as the epidemiological conditions deteriorated in many of those countries. As FDW11 explained: *“we have a lot of protocols, laws [in her home country], but the implementation is not that good*.*”*

Additionally, many FDWs reflected on the pressure they faced to send money back to their families. FDWs were often the primary breadwinners for their families prior to COVID-19, and the increasing financial difficulties in their home countries caused by the pandemic exacerbated the pressure they felt. As FDW9 said: *“For us the same salary … but our family … there’s no money and then we have to send more money to support them*.*”* Many FDWs became the sole earner of their family and sent most, if not all, of their salary back home. This practice was commonly framed as an obligation to their family, with FDW11 saying: *“As a first child I need to help my parents. I need to come here in Hong Kong to support my predecessors*.*”* KI1 also noticed that *“there is more demand on the finances”* from FDWs’ families.

Many FDW participants were greatly affected by the restrictions on international travel. As FDW12 said: *“I cannot go [to] my country, and I want [to] take care [of] my mom*.*”* Some had been unable to initially return to Hong Kong for work after Chinese New Year and many had friends who had lost jobs, which led to increased stress over their employment status. FDW1 recalled: *“So many of my friends already [got] terminated because they didn’t [come] back here in Hong Kong as schedule[d]*.*”* Others were unable to return to their home countries to see their families for their vacation, like FDW5 who said: *“I always go back home every July to take my vacation but unfortunately, I can’t do that this year because of the pandemic*.*”*

### 3.2 Changes in Work Situation

Almost all participants reported changes to their current work situation, including the actual work FDWs must perform for employers as well as their employment status.

#### 3.2.1 Employers’ treatment of FDWs

As the pandemic situation worsened in Hong Kong, many employers imposed additional restrictions on FDWs’ movements to try to minimise exposure to the virus. Based on a recommendation made by the government early into the pandemic, FDWs were encouraged to stay at home on their rest day. In many cases, however, employers forbade their FDW from leaving the house on their rest day. KI1 observed: *“The government has called for the domestic workers not to go out on Sunday … many employers just take this recommendation by the government as an instruction*.*”* Some FDW participants also experienced double standards in mobility restriction, where employers would do things that they forbade their FDWs from doing: *“[My employer is] always going out … they’re just afraid that I might bring home [the] virus … but they don’t think about themselves bringing the virus*.*”* (FDW7)

The nature of many FDWs’ work also changed during the pandemic, which made their job harder and put more pressure on them. Two FDW participants even described FDWs as *“front liners of the employers”* (FDW5 and FDW6), reflecting the essential nature of their work in the prevention and control of the virus at home. This caused many of them to feel stressed, tired, exhausted, and burned out. FDW9 stated: *“I work more, I clean more than usual. Because [I] have to be careful, everything must be cleaned*.*”* Furthermore, FDWs’ own worries about the pandemic situation in Hong Kong added pressure to keep the house clean. FDW10 said:

> *“Psychologically you really want to clean, clean, clean, clean. It’s very tiring but you really want to make sure, even though you know that it’s clean, but you still have to clean, clean, clean, because you’re afraid … Psychologically, it’s very draining*.*Physically it’s very tiring*.*”*

FDW participants also recalled instances where they or the people they knew were subjected to riskier working conditions. Due to the pandemic, many employers forced FDWs to clean with harsh chemicals, which caused adverse health effects. This has been noted by FDW7 and KI1, respectively:

> *“Sometimes it will make my head dizzy because the smell is too powerful, even [if] I use the mask, sometimes [it] make my chest hurt … sometimes I’m afraid that I might get sick, not just because of the COVID-19, but because of the chemical that I use in the everyday life*.*”*
>
> *“Some employers want … more saturated bleach to clean, and this affect[s] the workers’ [respiratory] health*.*”*

Some employers also put FDWs at an increased risk of COVID-19 exposure. For example, employers required to undergo quarantine at home did not provide alternative accommodation for FDWs during the 14-day period. KI1 shared: *“We have cases that worker’s employers, family, returned home and they don’t wear a mask [during quarantine]*.*”*

Finally, there were a few accounts of FDWs being abused by employers. KI1 stated: *“[FDWs have] less bargaining power [which keeps them silent] against the abuse and exploitation that they are facing*.*”* Some FDWs also decided to tolerate the abuse they faced in order to maintain their job security. As FDW11 put it: *“Sometimes [other FDWs] are too patient, because they don’t want to lose their job*.*”*

#### 3.2.2 Employment status of FDWs

Many FDWs expressed concerns about losing their jobs due to the pandemic. Some had heard stories of friends who had been terminated for COVID-related reasons, while others were worried about their employer’s ability to continue paying them, either because their employers had lost jobs themselves, or were leaving Hong Kong. FDW8 said: *“Many of the employers lost their jobs. So once he lost the job with his employers, that means migrant domestic workers will also be losing her job*.*”* A few FDW participants shared that they had recently renewed or planned to renew their contract with their current employer to ensure that they could keep working, especially given the uncertain nature of the pandemic. As FDW8 explained: *“Once [FDWs] lose our job we are thinking of course of our family back home, because that is the first or the very reason why we come here … not just for ourselves but for our family*.*”*

A few FDWs had also reported that they had actually lost their jobs due to the pandemic. As such, these FDW participants were under pressure to find new employment but expressed difficulties due to a lack of employment opportunities during the pandemic. Furthermore, there were a few accounts of unfair termination of FDWs. In some cases, employers would be worried about FDWs contracting COVID-19, especially when they showed flu-like symptoms. KI1 stated: *“You also should not discriminate when the domestic workers just had the flu … you cannot dismiss the workers*.*”* In other cases, FDWs would be terminated if they did not conform with employers’ restrictions. FDW5 shared an incident where an FDW she knew insisted to go out on her rest day, and *“when she come back her things were outside the house*.*”*

### 3.3 Hong Kong policies and blind spots

The FDW community was directly affected by the policies enacted by the Hong Kong government to control the spread of COVID-19, yet many of these policies and decisions were blind to the experiences of FDWs. Several FDWs felt that there was a lack of consideration for FDWs by policymakers, and that their needs were not being considered. Some expressed that they felt invisible in the eyes of the government, including FDW7 who said: *“We are just like nothing to them … we really need to work hard to make them hear us*.*”* This was also observed by KI1, who said that *“the government is not listening enough”* to FDWs. As FDW2 explained: *“[The Hong Kong government] care for their own people, they must also care for us … we also deserve help from them in times of this pandemic*.*”*

Several FDW participants also reported experiencing prejudiced attitudes and discrimination from the wider Hong Kong society. KI1 stated: *“There has already been discrimination faced by [FDWs], but this COVID-19 is revealing [a] more extreme and worsening situation of the discrimination*.*”* The most common stereotype cited by FDW participants was the perception of FDWs as potential carriers of COVID-19. FDW1 described this perception as: *“The virus is only active on Sundays [when it] will go out and then they will choose [foreign] domestic workers*.*”*

#### 3.3.1 Protection and support during the pandemic

Many FDWs felt that they lacked adequate protection and support from the Hong Kong government. Three key aspects of this were identified. First, there was a lack of allocation of quarantine sites by the government for FDWs. As FDW5 stated: *“We don’t have any houses [to] stay for quarantine but then we need to find [one] ourself*.*”* Many FDW participants reported difficulties finding a suitable place to complete their quarantine, and a few of those who did undergo quarantine did so under unsafe conditions. For example, one FDW participant completed her quarantine in her employer’s home while the family still lived there, while others did so in boarding house rooms with at least one other person. FDW11 recommended: *“The government should build a facility for foreign domestic workers, especially to support the accommodation*.*”*

Additionally, a lack of financial support from the government was reported. In June, the government announced the “Cash Payout Scheme,” which disbursed $10,000 HKD ($1,290 USD) to each eligible Hong Kong permanent resident aged 18 or above [38]. However, because FDWs are not entitled to become permanent residents of Hong Kong, none of them were eligible to benefit from this scheme.

As FDW2 put it: *“The local Hong Kongers get support from the government, but they also must include the workers because we are also affected by this virus*.*”*

FDW10 shared: *“My employer knows. We even talk about this … she’s also wondering why we are not given the subsidy*.*”*

Finally, there was a feeling of a lack of food and personal protective equipment (PPE) by the government. Many FDWs undergoing quarantine, or those who were unemployed, were unable to pay for food or PPE, and wished the government would provide them with these basic necessities. FDW6 said: *“Supposedly the government must provide us the mask, hand sanitizer or alcohol but it’s already been too late when they had given us those masks*.*”*

Perhaps due to the lack of support from the Hong Kong government, many FDW participants reported receiving support from other members of the FDW community and community organizations, most commonly in the form of financial support, particularly for those who were unemployed. FDW participants also reported sharing or receiving food and PPE, with food supplies being more likely to come from individual community members, and PPE most likely to come from community organizations such as labour unions. Furthermore, some participants also reported giving financial and emotional support to fellow FDWs, as well as sharing information about the virus. For example, FDW7 explained that she would *“translate to the others the information that we get from the English resource,”* and that other FDWs who understood Cantonese would do the same for those who could not understand either language.

## 4. Discussion

This study examined the effects of the COVID-19 pandemic on FDWs in Hong Kong. While several aspects of the FDW experience, such as job security, discrimination, and abuse, predate the pandemic, our findings suggest that the pandemic has further exposed many of these underlying conditions. It also highlights the challenges that many migrants faced during the pandemic, namely physical and emotional separation from family, discrimination by the host country, and the lack of inclusion of this population in state sponsored policies [10]. The COVID-19 pandemic has exacerbated inequalities and discrimination against FDWs, who are already constrained by a series of power relationships at the intersection of their race, gender, and socioeconomic status.

The finding that FDWs are experiencing the current pandemic across two countries seems to be a novel one, as we have found no previous studies that examined the effects of catastrophic events at home on overseas nationals. The diasporic experience of FDWs is not new, however, as it has previously been found that FDWs would continuously (re)negotiate their self-identity partly through maintaining connections to their families in their home countries [39]. By expanding on this finding, it is possible to consider how a global pandemic could affect their self-identity. Since many of their family members had been negatively impacted by the pandemic, their sense of identity in relation to their home countries seems to have strengthened – there may be more importance placed on their role as the breadwinner who can provide for their families’ increasing financial needs. However, worries about their families and home countries, which came up frequently during our interviews, may also be causing increased mental stress for FDWs. Additionally, the implications of this mounting pressure to provide for their families is that FDWs could be more willing to tolerate abusive employment relationships to continue sending remittances back home, amplifying their already vulnerable status. Further research is needed on the short- and long-term mental health effects of this dual-country experience, and how the FDW experience interacts with the home country experience of family members.

This study also found that the treatment of FDWs by employers has gotten worse during the pandemic, and they have continued to be neglected by the Hong Kong government, causing a disconnection between the FDWs’ experience of the acute effects of COVID-19 and blind spots in the policy response. Their low-status position indirectly comes with the expectation that they will be more flexible and willing to cooperate with changes, even if these changes may result in increased risks to their physical health. As front liners for their employers, FDWs are expected to endure more pressure from employers to protect the family from the virus, yet they are not given any support when doing so. Existing research shows that it is important to improve FDWs’ ability to access health information and make informed decisions about health-related issues, as they could be a key resource to break chains of infection within the community [40]. Many of the FDW participants in this study already reported seeking out health information on their own, so more efforts by relevant government departments to provide accurate and comprehensive information in ethnic minority languages is needed to support their search.

However, rather than acknowledging the important role of these essential workers in the city’s disease prevention activities and providing support for them, the government has instead chosen to further limit their opportunities for rest and social interaction with community members. In fact, the neglect of FDWs actually put the overall community at higher risk of infection by forcing FDWs to quarantine or spend more time in cramped and unsafe boarding houses, which were unsurprisingly linked to two important clusters of COVID-19. FDWs have repeatedly been urged by the government to comply with social distancing measures and stay home on their rest days, with no less than 15 press releases issued over the course of 2020 concerning the matter. This neglects the importance of FDWs’ rest days as an opportunity to physically separate themselves from their workplace, maintain social connections both in Hong Kong and in their home countries, or even engage in activism or volunteer work. By continuing to draw attention to FDWs for their supposed lack of compliance, this suggests that the public should be particularly concerned about FDWs, and may have contributed to the discrimination, double standards and employer-imposed restrictions mentioned by our participants.

Several key issues emerged that highlighted the need for changes in policies related to FDWs in Hong Kong. Specific recommendations made by our FDW participants included more financial support, better access to food and PPE, and quarantine arrangements provided by the government. Like other members of Hong Kong society [35], FDWs in our study reported turning to community support networks for sharing PPE and other essentials during the pandemic. However, many also felt that they did not have the power to speak up to the government and demand change, illustrating their low status and lack of power in Hong Kong society.

The situation of FDWs in Hong Kong at a policy level has seen some improvement since the end of our data collection period. In light of the COVID-19 clusters linked to temporary boarding houses, the Hong Kong government offered free, one-off COVID-19 tests for FDWs staying in such facilities in August [41]; this was offered once again in December and extended to all FDWs in Hong Kong [42]. On November 3, the government reminded employers not to dismiss FDWs who had contracted COVID-19, as it would be in violation of the Disability Discrimination Ordinance [43]. In January 2021, the Ombudsman announced that it would launch an investigation into boarding house conditions and the government’s role in the regulation of such facilities [44]. However, there are still many problems that need to be addressed. For example, it was announced on December 4 that fixed penalties for those who violated anti-epidemic measures would increase from $2,000 HKD to $5,000 HKD ($258 USD to $645 USD) [45], with a separate press release on the same day reminding FDWs in particular of this change. However, given that this is more than an FDW’s minimum salary, this could create a heavy financial burden on those who get fined. FDWs continue to be reminded to comply with social distancing measures, and are highlighted in the media when they do not do so. Although progress is being made, there is clearly still much to be done to ensure FDWs are considered and supported in Hong Kong’s COVID-19 response.

### 4.1 Limitations

This study has several important limitations. FDWs were recruited as part of a larger project on gender and thus had not been the sole focus of the study, so our interview guide was not tailored specific to the concerns of FDWs. The views expressed by FDW participants in this study may not fully represent the experiences of all FDWs working in Hong Kong or generalize to the pandemic experiences of FDWs in other countries. Participants were recruited through social media adverts, so those who chose to respond many have had a vested interest in sharing their experiences and opinions on this topic. As all interviews were conducted in English, this may have limited the amount of information participants were able to share as non-native speakers and excluded those who could not speak the language.

Future research is needed that is specifically catered to the experiences of FDWs, including comparisons with FDWs in other countries. Given this, future studies should also recruit larger and more robust sample sizes to increase generalizability of study findings. Since there were many suggestions for change that came up during our interviews, future research could consult with FDW collectives who have perhaps attempted to gain recognition from the government to explore how the COVID-19 pandemic may be used as a basis to claim greater recognition and rights for FDWs. Given the ever-evolving nature of this pandemic, future studies will be needed that can measure what has gotten worse or better since the start of the pandemic. The use of a mixed methods design may enable future researchers to increase the depth and breadth of study findings.

## 5. Conclusions

The current study demonstrates that the COVID-19 pandemic has exacerbated the existing power dynamics that constrain FDWs in Hong Kong. Not only is there a greater need to provide for family members back in FDWs’ home countries, but there are also increased pressures from employers and a lack of support from the Hong Kong government. Through this study’s findings of the current situation and challenges faced by FDWs in Hong Kong during the COVID-19 pandemic, it is clear that policy-level interventions are needed to mitigate the particularly negative effects on FDWs. More supportive policies should be adopted that not only consider the specific needs of FDWs but listens to them.

## Data Availability

Due to the nature of this research, participants of this study did not agree for their data to be shared publicly, so supporting data is not available.

## Acknowledgements

We thank the FDW participants for their trust in sharing their stories with us. We thank Ka Chun Yung for his help in creating the initial coding guide.

## Footnotes

1 More details about this project can be found at http://www.genderandcovid-19.org

